# Communicating about COVID-19 vaccine development and safety

**DOI:** 10.1101/2021.06.25.21259519

**Authors:** Alistair Thorpe, Angela Fagerlin, Jorie Butler, Vanessa Stevens, Frank A. Drews, Holly Shoemaker, Marian Riddoch, Laura D. Scherer

**Author notes:** **Corresponding author**: Alistair Thorpe PhD; University of Utah School of Medicine, Department of Population Health Sciences, 295 Chipeta Way, Williams Building, Room 1N410, Salt Lake City, Utah 84108.

## Abstract

**Purpose:** Beliefs that the risks from the vaccine outweigh the risks from getting COVID-19 and concerns that the vaccine development process was rushed and lacking rigor have been identified as important drivers of hesitancy and refusal to get a COVID-19 vaccine. We tested whether messages designed to address these beliefs and concerns might promote intentions to get a COVID-19 vaccine.

**Method:** An online survey fielded between March 8-March 23, 2021 with US Veteran (*n*=688) and non-Veteran (*n*=387) respondents. In a between-subjects experiment, respondents were randomly assigned to a control group (with no message) or to read one of two intervention messages: 1. a fact-box styled message comparing the risks of getting COVID-19 compared to the vaccine, and 2. a timeline styled message describing the development process of the COVID-19 mRNA vaccines.

**Results:** Most respondents (60%) wanted a COVID-19 vaccine. However, 17% expressed hesitancy and 23% did not want to get a COVID-19 vaccine. The fact-box styled message and the timeline message did not significantly improve vaccination intentions, *F*(2,358)=0.86, *p*=.425, 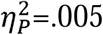, or reduce the time respondents wanted to wait before getting vaccinated, *F*(2,306)=0.79, *p*=.453, 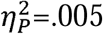, compared to no messages.

**Discussion:** We did not find an impact on vaccine intention based on providing information about vaccine risks and development. Further research is needed to identify how to effectively address concerns about the risks associated with COVID-19 vaccines and the development process and to understand additional factors that influence vaccine intentions.

## Introduction

All of the currently available COVID-19 vaccines have demonstrated high efficacy against COVID-19 (Olliaro, 2021). Maximizing the public health benefits from these vaccines depends on achieving high levels of vaccine coverage. One of the major barriers to achieving widespread coverage of COVID-19 vaccines is public hesitancy and reluctance to receive them (Sallam, 2021). Prior research has established that attitudes towards vaccines and intentions to receive or refuse them are driven by a multitude of factors (Larson et al., 2014). Within this literature, there is consistent evidence that people who are hesitant or reluctant to receive a vaccine often have doubts about the benefits of vaccines and concerns about the general safety of taking them (Karafillakis & Larson, 2017).

Throughout the pandemic, how people perceive the threat posed by COVID-19 has been shown to vary based on a number of factors such as age (Bruine de Bruin, 2020), political ideology (Calvillo et al., 2020; Kerr et al., 2021), and philosophical beliefs (Byrd & Białek, 2021). In extreme cases, some people believe that COVID-19 does not pose a threat at all as they question the legitimacy of the pandemic and deny the existence of the virus (Gorski, 2020). How people perceive the risk of COVID-19 matters as those who do not perceive COVID-19 to be a serious threat are likely to undervalue the benefits of vaccination (Karafillakis & Larson, 2017) and are therefore less likely to want to receive a vaccine (Betsch, et al., 2018; Malik et al., 2020). Furthermore, people who do not consider COVID-19 to present a serious threat may also be reluctant to receive a vaccine because they are more likely to perceive that they are more at risk from potential harms from vaccination than from acquiring COVID-19 (Karafillakis & Larson, 2017; Betsch, et al., 2018).

The perceived safety of a vaccine is another factor that can have a strong influence on peoples’ decisions about whether or not to get vaccinated (Karafillakis & Larson, 2017; Karlsson et al., 2021). Current evidence suggests that people who are hesitant about receiving a COVID-19 vaccine are often concerned about the safety of the COVID-19 vaccine development process (Dror et al., 2020). Producing a number of highly effective vaccines within a year of an infectious disease outbreak is undoubtedly one of humanity’s greatest medical achievements. It is important to acknowledge that this achievement was only possible because of decades of prior research and clinical trials on vaccine technology (e.g., mRNA and viral vectors) and previous coronavirus outbreaks (e.g., SARS and MERS) (Ball, 2020; Li et al., 2020). However, the importance and rigor of this prior research has often been overlooked in public discourse, which has more commonly emphasized the speed of development. As a consequence, concerns that the safety of the vaccines was compromised by rushed development and the use of experimental or untested technology have become widespread (Dror et al., 2020). High-profile media coverage of very rare side-effects following COVID-19 vaccination (e.g., anaphylaxis and thrombosis with thrombocytopenia syndrome) may have compounded this issue by disproportionately raising public concern about vaccine safety and further fueling public hesitancy (Dodd et al., 2021; Tran et al., 2018). In addition, it is also possible that the emergency use authorization (EUA) given to the COVID-19 vaccines in response to the severity of the COVID-19 pandemic also resulted in concerns about the safety of the vaccines and, in turn, increased public hesitancy (Kreps et al., 2020; Quinn et al., 2009).

In summary, people who are hesitant about receiving a COVID-19 vaccine may believe that the risks from the vaccine outweigh the risks from getting COVID-19 or have concerns about the safety of the vaccine development process. Developing and testing the efficacy of communications designed to directly address frequently cited concerns about the COVID-19 vaccines represents an important goal for research. Particularly as many government and health organizations aspire to provide this type of reassuring information, typically on their websites (e.g., https://www.cdc.gov/coronavirus/2019-ncov/vaccines/safety/safety-of-vaccines.html) and social media accounts. In the present study we tested whether messaging about the comparative risks of getting COVID-19 vs. the risk of COVID-19 vaccines, and also messaging about the timeline of mRNA vaccine development, might promote intentions to get a COVID-19 vaccine.

## Materials and method

### Study population and recruitment

Respondents were recruited by Qualtrics Online Panels between March 8-March 23, 2021. This experiment was conducted in the third wave of a three-wave longitudinal study. The study was administered online (in English) and was approved by the relevant IRBs. Respondents were compensated for their participation based on the terms of their panel agreement.

A total of 1075 respondents completed the third wave of the study, of which 688 (64%) were United States Veterans; median age of the sample was between 55 and 74 years old; 841 respondents (78%) were male, 819 respondents (76%) were non-Hispanic White, and the median household income was $70,000-$99,999. Sample retention from the first wave was 52% overall, 65% for Veteran and 38% for non-Veterans. There were 361 (33%) respondents who reported having not received any vaccine doses, 243 (23%) who reported having received 1 dose, and 471 (44%) who reported having received 2 doses. Respondents who reported having received at least 1 dose of a COVID-19 vaccine were not included in the experimental component of the survey, leaving a final sample of 361 respondents (Table 1).

**Table 1.** Respondent demographics.

|  | N (%) |
| --- | --- |
| *Age* |  |
| 18 to 34 | 46 (13%) |
| 35 to 54 | 80 (22%) |
| 55 to 74 | 205 (57%) |
| 75 or older | 28 (8%) |
| Did not respond | 2 (<1%) |
| *Gender* |  |
| Female | 123 (34%) |
| Male | 235 (65%) |
| Non-binary/Third gender or Transgender man/Transman | 3 (1%) |
| *Race/Ethnicity* |  |
| Non-Hispanic White | 252 (70%) |
| Non-Hispanic Black | 49 (14%) |
| Hispanic | 32 (9%) |
| Asian/Asian American | 15 (4%) |
| American Indian/Alaskan Native | 2 (1%) |
| Another race | 7 (2%) |
| Multiracial | 4 (1%) |
| *Income* |  |
| \$0 - \$49k | 131 (36%) |
| \$50K to \$99K | 125 (35%) |
| \$100K or more | 87 (24%) |
| Prefer not to say | 18 (5%) |
| *Residence* |  |
| Rural | 77 (21%) |
| Small (less than 100,000) | 58 (16%) |
| Suburban near large city | 157 (44%) |
| Mid-sized city<br>(100,000 to 1million) | 29 (8%) |
| large city more than 1 million | 40 (11%) |

### Procedure

In a fully between-subjects experiment, respondents saw either a Schwartz et al. (2007) factbox style message about the risks of getting a vaccine compared to getting COVID-19 (factbox message), a message about the development of COVID-19 vaccines (timeline message; both messages are presented in Figure 1), or no message (control group). Respondent assignment to study conditions was carried out using the built-in randomizer function in Qualtrics’ survey flow.

**Figure 1.**
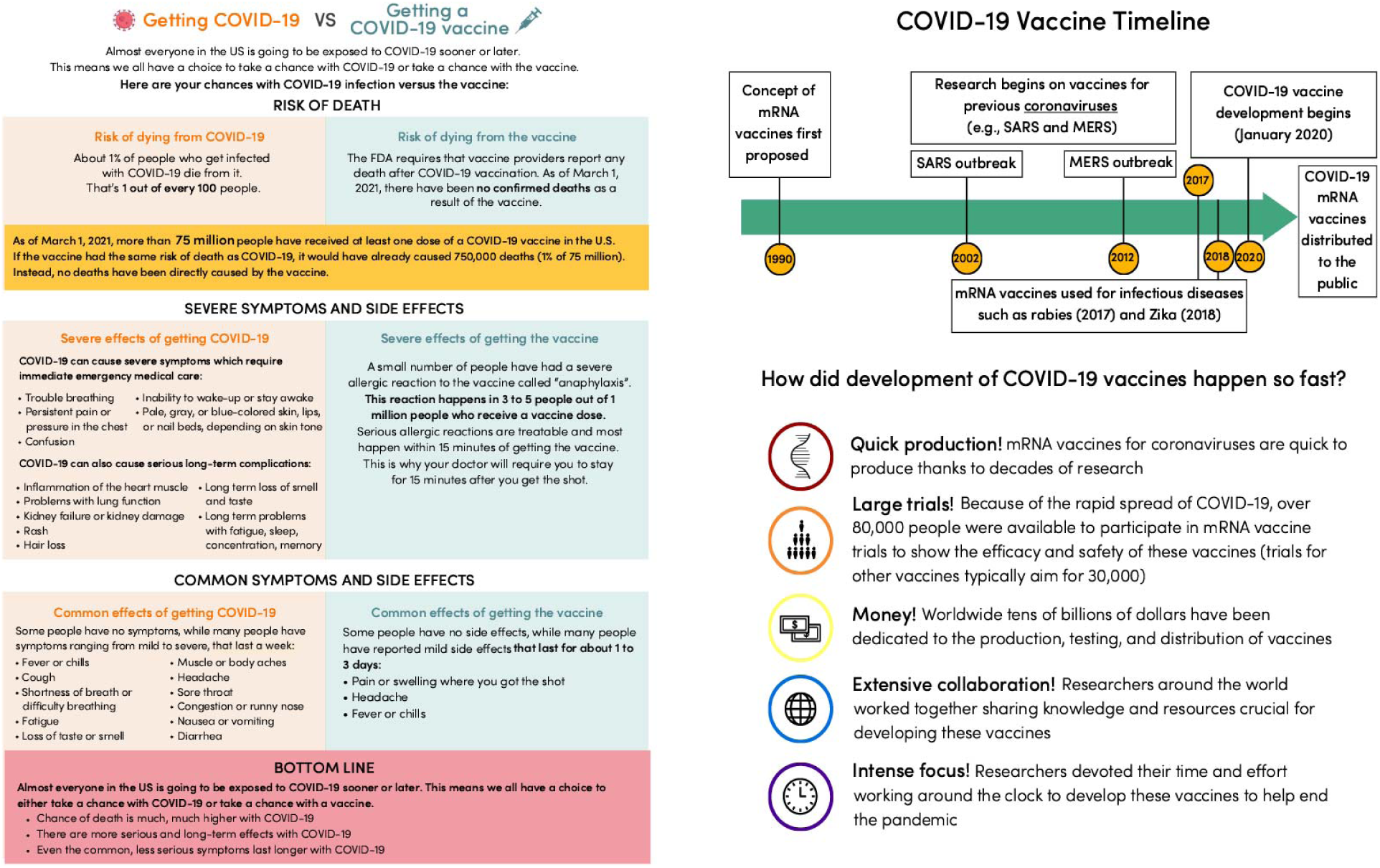
Factbox style message (left) about the risks of getting a vaccine compared to getting COVID-19 and the Timeline message (right) about the development of COVID-19 vaccines.

Respondents then reported their COVID-19 vaccine intentions (“When a coronavirus vaccine becomes available to you, how interested are you in getting the vaccine?”) using a 5-point scale (1=“I definitely do NOT want the vaccine”, 2= “I do NOT want to get the vaccine”, 3=“Unsure”, 4=“I WANT to get the vaccine”, 5=“I definitely WANT the vaccine”) and how long they intended to wait before getting vaccinated (“How soon after the COVID-19 vaccine becomes available to you would you become vaccinated?”) using an 8-point scale (1=“Immediately”, 2=“Less than one month”, 3=“One month to less than 3 months”, 4=“3 months to less than six months”, 5=“6 months to less than 1 year”, 6=“1 year to less than 2 two years”, 7=“I would wait 2 years or more”, 8=“I would never get it”).

We also asked respondents about their views regarding COVID-19 vaccine safety (“In your view, how safe is the COVID-19 vaccine?”) using a 5-point scale (1=“Not at all safe”, 2= “Slightly safe”, 3=“Somewhat safe”, 4=“Moderately safe”, 5=“Extremely safe”) and the possibility of experiencing side effects (“How worried are you about experiencing side effects from the COVID-19 vaccine?” using a 5-point scale (1=“Not at all concerned”, 2= “Slightly concerned”, 3=“Somewhat concerned”, 4=“Moderately concerned”, 5=“Extremely concerned”).

### Pre-registered analyses

We ran two omnibus one-way ANOVA analyses to examine whether the COVID-19 vaccine risk comparison factbox message and vaccine development timeline message influenced 1) respondents’ intentions to get vaccinated and 2) the time they would wait to get vaccinated. Analyses were repeated controlling for age and gender.

We ran two additional omnibus one-way ANOVAs to examine whether 1) the factbox message comparing the risks associated with getting COVID-19 to the risks associated with receiving a COVID-19 vaccine reduced respondents’ concern about experiencing side effects from the COVID-19 vaccine and 2) whether the timeline message regarding the vaccine development process increased respondents’ views about how safe the COVID-19 vaccine is. All analyses were performed using RStudio statistical software (RStudio Team, 2021).

## Results

Contrary to our expectations, we found no differences between groups regarding vaccination intentions, *F*(2,358)=0.86, *p*=.425, 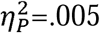, or the time respondents would wait to get vaccinated, *F*(2,306)=0.79, *p*=.453, 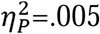. In fact, as shown in Figure 2, mean scores for both the intervention groups suggest that respondents were slightly *less* willing to get a COVID-19 vaccine and also would want to wait longer before getting one compared to those in the control group who did not see any message.

**Figure 2.**
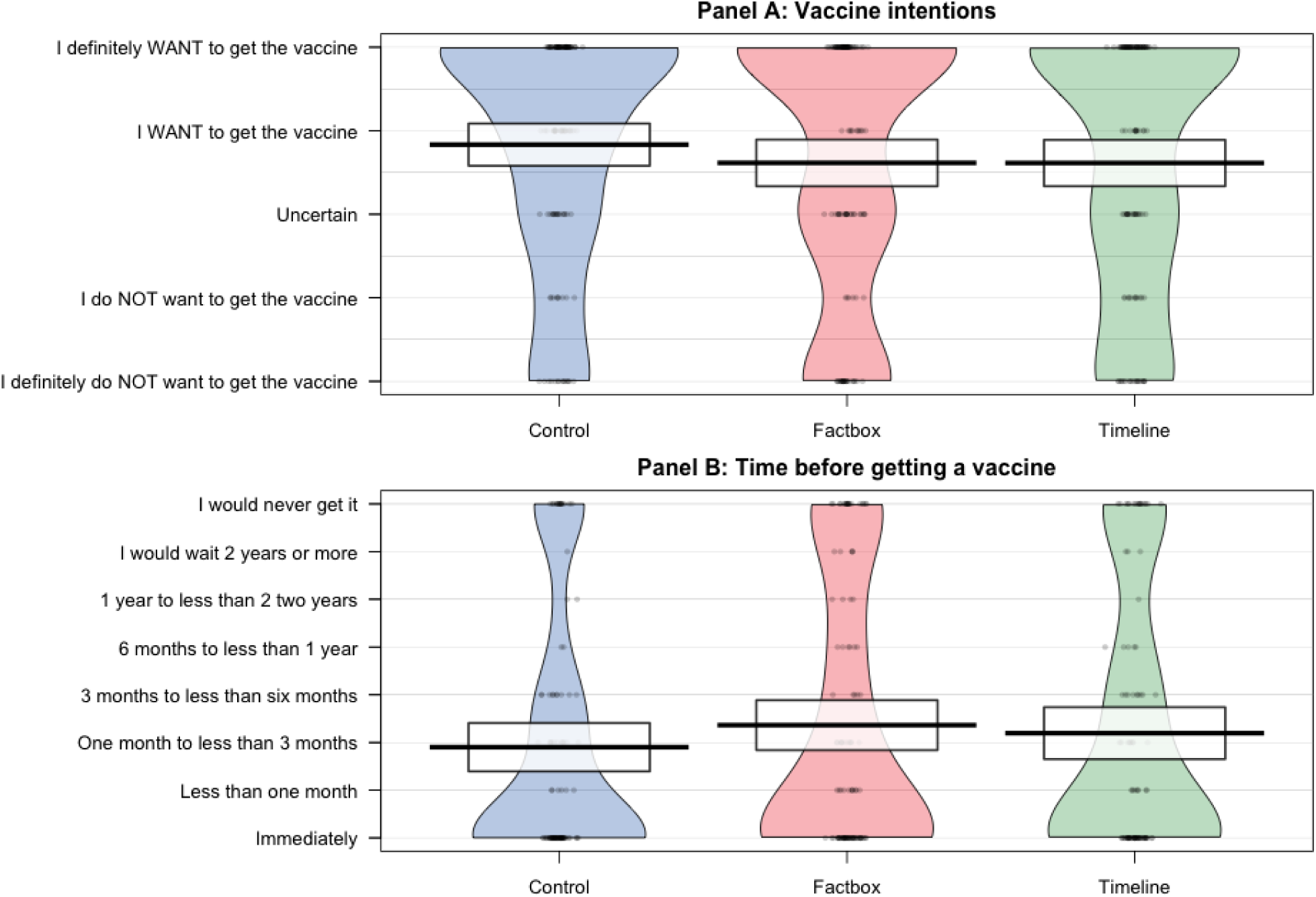
Mean COVID-19 vaccine intention scores for the factbox message (showing risk of getting COVID-19 versus risk of getting a COVID-19 vaccine), the timeline message (showing the development of mRNA and COVID-19 vaccine research), and control group. The middle bold line represents the mean and the box borders represent 95% confidence intervals. Individual data points are displayed within the shaded density distributions.

These results did not differ when controlling for respondents’ age, (no effect of group on vaccination intentions, *F*(2,355)=0.06, *p*=.945, 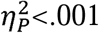, and no effect of group on time respondents would wait to get vaccinated, *F*(2,303)=0.05, *p*=.950, 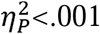, or gender, (no effect of group on vaccination intentions, *F*(2,352)=1.46, *p*=.233, 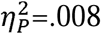, and no effect of group on time respondents would wait to get vaccinated, *F*(2,301)=1.54, *p*=.217, 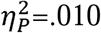. Furthermore, views about how safe COVID-19 vaccines are, *F*(2,358)=0.20, *p*=.815, 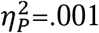, and worry about side effects, *F*(2,357)=1.89, *p*=.152, 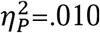, did not differ between groups. Again, these findings were contrary to our initial prediction.

### Exploratory analyses and results

We conducted exploratory analyses using omnibus one-way ANOVAs to examine the influence of the messages on vaccine intentions and time to getting vaccinated on two subsets of the total sample. These analyses were not planned a-priori, but were conducted after the unexpected finding that neither of the messages had an impact on respondents’ vaccination intentions or the time they wanted to wait before getting vaccinated.

To examine the possibility that any impact of the messages on vaccination intentions could be dependent on whether respondents took the time to read and process the information provided in them, we re-ran our analyses on a subset of respondents who spent at least 10 seconds on the page displaying the intervention messages (n=298). We found that results from this subset of respondents did not differ from the overall sample: there were no significant differences in vaccination intentions, *F*(2,295)=1.51, *p*=.224, 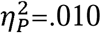, and no significant difference in time respondents would wait to get vaccinated between groups, *F*(2,252)=1.08, *p*=.342, 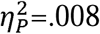.

Another possibility is that any impact of the messages on vaccination intentions may be limited only to respondents who were initially hesitant about getting a COVID-19 vaccine (Freeman, et al. 2021). That is, an effect of the intervention might be dampened by ceiling effects on vaccination intentions. For the second subset we only selected respondents who had reported that they were hesitant about getting a COVID-19 vaccine in an earlier survey fielded in December 2020 (i.e., reported that they either “do NOT want to get the vaccine” or “definitely do NOT want to get the vaccine”). Although this subset included a small number of participants, there were again no significant differences between groups on respondents’ vaccination intentions, *F*(2,83)=0.32, *p*=.724, 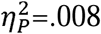, or the time they would wait to get vaccinated, *F*(2,61)=1.05, *p*=.356, 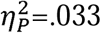.

## Discussion

Major advances in vaccine development are at risk of being undermined by increasing levels of vaccine hesitancy (WHO, 2019). In the immediate term, public hesitancy towards COVID-19 vaccines presents a notable barrier to stopping the spread of COVID-19 and ending the pandemic (Sallam, 2021). Messaging designed to address prominent concerns about COVID-19 vaccines might encourage people who are otherwise hesitant about getting vaccinated to receive one.

In the present study we found no evidence that the messages comparing the potential risks of getting COVID-19 to the potential risks associated with COVID-19 vaccines and regarding the vaccine development process, had any effect on respondents’ intentions to get a COVID-19 vaccine, the time that they would want to wait before getting vaccinated, perceived safety of COVID-19 vaccines, or worry about vaccine-related side effects. These results were unanticipated and remained consistent when controlling for the age and gender of respondents and following exploratory analyses of respondents who spent at least 10 seconds looking at the messages and those who had previously expressed that they did not want to get a COVID-19 vaccine. The messages were designed to address concerns about the risks from COVID-19 vaccines and the vaccine development process as they are reasons frequently cited by people who are hesitant about receiving a COVID-19 vaccine (Dodd et al., 2021; Dror et al., 2020). The present findings are unfortunate as effective low-cost messaging strategies could contribute substantially to combatting growing levels of vaccine hesitancy and help end the COVID-19 pandemic.

One potential reason for these null findings could simply be that the information in the two messages was not sufficiently convincing to influence the vaccine intentions of people who are concerned about the risks of vaccines and the development process. For individuals who did not have these concerns, it could also have been the case that the information in the messages may have drawn their attention to these issues which may have then served to undermine their prior confidence in vaccine development and safety (Betsch et al., 2011). It is unlikely that these null effects were due to insufficient statistical power. Observed effect sizes in this experiment were very small (such that even if significant, the meaningfulness of these differences would have been questionable), and if anything, the intervention groups showed slightly more vaccine hesitancy than the control group, not less.

Given the cross-sectional nature of this experiment, it is not possible to examine whether repeated exposure to these messages may have yielded more positive results. Repeated messaging about health issues is often more effective at changing attitudes and behaviors (Frew, et al., 2014), however, given the prevalence of media coverage about COVID-19 and vaccine development it is also possible that efficacy of these messages may be suppressed by respondents having high prior exposure to similar information.

It is also possible that the design of the message–regarding both the style and content, may have also contributed to the null finding. However, given the data available from the present study it is not possible to posit informed justifications for the null findings based on respondents’ perceptions of the messages themselves. This does prompt an avenue for future research, which might leverage qualitative methods (e.g., focus groups or open responses) in order to better understand how people perceived the messages used in the present study.

The present findings are aligned with research which suggests that providing corrective information based on an information-deficit assumption is not always effective at addressing people’s concerns and doubts (Dubé et al., 2015; Nyhan et al., 2014; Nyhan & Reifler, 2015). The present findings also highlight the need for testing public health messages to ensure they are as effective as possible. Future research might explore whether different message styles or sources (e.g., non-partisan experts or popular figures) may be more effective at sufficiently addressing concerns about vaccine development and safety. We also believe these findings are important in highlighting the need for research into alternative influences of vaccine intentions that might be more receptive to messaging than concerns about safety and development.

### Limitations

Despite evidence that self-reports are good predictors of health behaviors (Webb & Sheeran, 2006), it is important that these findings are considered in the context of known limitations of this method (e.g., social desirability). The study design (i.e., US-based recruitment, online, and in English) prevents generalization of the present findings outside of the US and to people who may have limited internet access and lower English proficiency. Furthermore, our sample consists of both Veteran and non-Veteran respondents completing the third (and final) survey of a longitudinal study, which began in December, 2020 and thus are not representative of the general population.

## Conclusion

In the present study, providing corrective information about the risks of getting COVID-19 compared to receiving a COVID-19 vaccine and about the development of COVID-19 vaccines was not effective at promoting intentions to get a COVID-19 vaccine. Further research is needed to identify how to effectively address concerns about the risks of side effects from COVID-19 vaccines and the vaccine development process.

## Data Availability

Data and study materials are available from the first author upon request.

## Acknowledgments

The views expressed in this paper are those of the authors and do not necessarily represent the position or policy of the U.S. Department of Veterans Affairs or the United States Government. Data and study materials are available from the first author upon request. The pre-registration protocol for this study is available at: https://aspredicted.org/blind.php?x=az6k8w

## References

Ball, P. (2020). The lightning-fast quest for COVID vaccines—And what it means for other diseases. Nature, 589(7840), 16–18. https://doi.org/10.1038/d41586-020-03626-1

Betsch, C., Ulshöfer, C., Renkewitz, F., & Betsch, T. (2011). The Influence of Narrative v. Statistical Information on Perceiving Vaccination Risks. Medical Decision Making, 31(5), 742–753. https://doi.org/10.1177/0272989X11400419

Betsch, C., Schmid, P., Heinemeier, D., Korn, L., Holtmann, C., & Böhm, R. (2018). Beyond confidence: Development of a measure assessing the 5C psychological antecedents of vaccination. PLoS ONE, 13 (12), 1-32. 10.1371/journal.pone.0208601

Bruine de Bruin, W. (2020). Age Differences in COVID-19 Risk Perceptions and Mental Health: Evidence From a National U.S. Survey Conducted in March 2020. The Journals of Gerontology Series B: Psychological Sciences and Social Sciences. https://doi.org/10.1093/geronb/gbaa074

Byrd, N., & Białek, M. (2021). Your health vs. my liberty: Philosophical beliefs dominated reflection and identifiable victim effects when predicting public health recommendation compliance during the COVID-19 pandemic. Cognition, 212, 104649. https://doi.org/10.1016/j.cognition.2021.104649

Calvillo, D. P., Ross, B. J., Garcia, R. J. B., Smelter, T. J., & Rutchick, A. M. (2020). Political Ideology Predicts Perceptions of the Threat of COVID-19 (and Susceptibility to Fake News About It). Social Psychological and Personality Science, 11(8), 1119–1128. https://doi.org/10.1177/1948550620940539

Dodd, R. H., Pickles, K., Nickel, B., Cvejic, E., Ayre, J., Batcup, C., Bonner, C., Copp, T., Cornell, S., Dakin, T., Isautier, J., & McCaffery, K. J. (2021). Concerns and motivations about COVID-19 vaccination. The Lancet. Infectious Diseases, 21(2), 161–163. https://doi.org/10.1016/S1473-3099(20)30926-9

Dror, A. A., Eisenbach, N., Taiber, S., Morozov, N. G., Mizrachi, M., Zigron, A., Srouji, S., & Sela, E. (2020). Vaccine hesitancy: The next challenge in the fight against COVID-19. European Journal of Epidemiology, 35(8), 775–779. https://doi.org/10.1007/s10654-020-00671-y

Dubé, E., Gagnon, D., & MacDonald, N. E. (2015). Strategies intended to address vaccine hesitancy: Review of published reviews. Vaccine, 33(34), 4191–4203. https://doi.org/10.1016/j.vaccine.2015.04.041

Freeman, D., Loe, B. S., Yu, L. M., Freeman, J., Chadwick, A., Vaccari, C., … & Lambe, S. (2021). Effects of different types of written vaccination information on COVID-19 vaccine hesitancy in the UK (OCEANS-III): a single-blind, parallel-group, randomised controlled trial. The Lancet Public Health. doi:10.1016/S2468-2667(21)00096-7

Frew, P. M., Owens, L. E., Saint-Victor, D. S., Benedict, S., Zhang, S., & Omer, S. B. (2014). Factors associated with maternal influenza immunization decision-making: Evidence of immunization history and message framing effects. Human vaccines & immunotherapeutics, 10(9), 2576–2583. https://doi.org/10.4161/hv.32248

Gorski, D. (2020, April 20). COVID-19 pandemic deniers and the antivaccine movement: An unholy alliance. Science-Based Medicine. https://sciencebasedmedicine.org/covid-19-pandemic-deniers-and-the-antivaccine-movement-an-unholy-alliance/

Karafillakis, E., & Larson, H. J. (2017). The benefit of the doubt or doubts over benefits? A systematic literature review of perceived risks of vaccines in European populations. Vaccine, 35(37), 4840–4850. https://doi.org/10.1016/j.vaccine.2017.07.061

Kerr, J., Panagopoulos, C., & van der Linden, S. (2021). Political polarization on COVID-19 pandemic response in the United States. Personality and Individual Differences, 179, 110892. https://doi.org/10.1016/j.paid.2021.110892

Kreps, S., Prasad, S., Brownstein, J. S., Hswen, Y., Garibaldi, B. T., Zhang, B., & Kriner, D. L. (2020). Factors Associated With US Adults’ Likelihood of Accepting COVID-19 Vaccination. JAMA Network Open, 3(10), e2025594. https://doi.org/10.1001/jamanetworkopen.2020.25594

Larson, H. J., Jarrett, C., Eckersberger, E., Smith, D. M. D., & Paterson, P. (2014). Understanding vaccine hesitancy around vaccines and vaccination from a global perspective: A systematic review of published literature, 2007–2012. Vaccine, 32(19), 2150–2159. https://doi.org/10.1016/j.vaccine.2014.01.081

Li, Y.-D., Chi, W.-Y., Su, J.-H., Ferrall, L., Hung, C.-F., & Wu, T.-C. (2020). Coronavirus vaccine development: From SARS and MERS to COVID-19. Journal of Biomedical Science, 27(1), 104. https://doi.org/10.1186/s12929-020-00695-2

Malik, A. A., McFadden, S. M., Elharake, J., & Omer, S. B. (2020). Determinants of COVID-19 vaccine acceptance in the US. EClinicalMedicine, 26, 100495. https://doi.org/10.1016/j.eclinm.2020.100495

Nyhan, B., & Reifler, J. (2015). Does correcting myths about the flu vaccine work? An experimental evaluation of the effects of corrective information. Vaccine, 33(3), 459–464. https://doi.org/10.1016/j.vaccine.2014.11.017

Nyhan, B., Reifler, J., Richey, S., & Freed, G. L. (2014). Effective Messages in Vaccine Promotion: A Randomized Trial. Pediatrics, 133(4), e835–e842. https://doi.org/10.1542/peds.2013-2365

Olliaro, P. (2021). What does 95% COVID-19 vaccine efficacy really mean? The Lancet Infectious Diseases, 0(0). https://doi.org/10.1016/S1473-3099(21)00075-X

Quinn, S. C., Kumar, S., Freimuth, V. S., Kidwell, K., & Musa, D. (2009). Public Willingness to Take a Vaccine or Drug Under Emergency Use Authorization during the 2009 H1N1 Pandemic. Biosecurity and Bioterrorism: Biodefense Strategy, Practice, and Science, 7(3), 275–290. https://doi.org/10.1089/bsp.2009.0041

RStudio Team. (2021). RStudio: Integrated Development Environment for R. RStudio. [RStudio]. PBC, Boston, MA. http://www.rstudio.com/.

Sallam, M. (2021). COVID-19 vaccine hesitancy worldwide: A concise systematic review of vaccine acceptance rates. Vaccines, 9(2), 160.

Schwartz, L. M., Woloshin, S., & Welch, H. G. (2007). The Drug Facts Box: Providing Consumers with Simple Tabular Data on Drug Benefit and Harm. Medical Decision Making, 27(5), 655–662. https://doi.org/10.1177/0272989X07306786

Tran, B. X., Boggiano, V. L., Nguyen, L. H., Latkin, C. A., Nguyen, H. L. T., Tran, T. T., Le, H. T., Vu, T. T. M., Ho, C. S., & Ho, R. C. (2018). Media representation of vaccine side effects and its impact on utilization of vaccination services in Vietnam. Patient Preference and Adherence, 12, 1717–1728. https://doi.org/10.2147/PPA.S171362

Webb, T. L., & Sheeran, P. (2006). Does changing behavioral intentions engender behavior change? A meta-analysis of the experimental evidence. Psychological Bulletin, 132(2), 249–268. https://doi.org/10.1037/0033-2909.132.2.249

WHO. (2019). Vaccination: European Commission and World Health Organization join forces to promote the benefits of vaccines. https://www.who.int/news/item/12-09-2019-vaccination-european-commission-and-world-health-organization-join-forces-to-promote-the-benefits-of-vaccines

